# Cytokines (IL-17, IL-23 and IL-33) in Systemic lupus erythematosus in Trinidad and Tobago

**DOI:** 10.1101/2020.09.27.20202762

**Authors:** Angel Justiz Vaillant, Patrick E Akpaka

## Abstract

Systemic lupus erythematosus (SLE) is the most common autoimmune disease. It is characterized by the presence of hundreds of autoantibodies against many organs and tissues, including the presence of a large number of autoantibodies, which are specific to self-antigens mainly of nuclear origin such as Smith antigen, double-stranded DNA (dsDNA), anti-Sjögren’s syndrome-related antigen A and B (SSA/Ro and SSB/La, respectively) and ribonucleoproteins, which are the hallmarks of the disease. Type I and II interferons, interleukin-6 (IL-6), IL-1, tumor necrosis factor-alpha (TNF-*α*), and immunomodulatory cytokines such as IL-10 and TGF-*β* are essential players in SLE. Additionally, T-cell-derived cytokines such as IL-17, IL-21, and IL-2 are dysregulated in SLE. In this study among cohorts of 60 individuals attending the hospital clinics in Trinidad and Tobago, blood samples were analyzed and the levels of the essential cytokines were measured using SLE Disease Activity Index (SLEDAI) 2000 score. The results confirmed that serum IL-17 and IL-23 levels were positively correlated with the SLE Disease Activity Index (SLEDAI) 2000 score in these patients. These findings have diagnostic and therapeutic implications. However, more work must be done targeting other cytokines relevant to autoimmunity and SLE in particular. Interleulin-33 is not an SLE marker, as has been noted in other populations.

## Introduction

Studies have provided new information on interleukins 17, 23, and 33 and systemic lupus erythematosus (SLE). In their study, Wong et al., 2008 showed that the production of IL-17 was uncharacteristically high in SLE patients, and it showed a positive correlation between IL-17 and the SLEDAI score [1]. Fabien et al., in 2013, showed that IL-17 serum levels were higher in patients with lupus than controls. However, these interleukin levels did not correlate with the SLE Disease Activity Index (SLEDAI) 2000 score [2]. A previous study also found that the association between IL-23 and the IL-17 axis is vital for the inflammatory response in SLE [1]. In a study conducted by Mok et al. (2010), their investigations showed no correlation was found between serum IL-17 and 23 in SLE patients with SLE. This study also showed higher serum IL-17 and IL-23 levels, and the lack of correlation between these cytokines suggests independent regulatory mechanisms [3]. A study in 2011 by Yang et al. found that IL-33 does not play a part in the illness but is needed in the acute phase and affects red blood cells and platelets [4]. The rationale for our study was the measurements of IL-17, IL-23, and IL-33 in the serum of patients suffering from systemic lupus erythematosus in Trinidad and Tobago; studying and determining these cytokines will be a tremendous diagnostic means and in further management of SLE patients in the country and the region.

## Materials and Methods

### Population, study design, and setting

All the experiments were conducted in accordance with the Declaration of Helsinki (1964). This study describes experimental work that carries a risk of harm to human subjects, and it was conducted with the human subjects’ understanding and consent, as well as a statement that the Ethical Committee of the University of the West Indies has approved the experiments.

A previous research project with fewer subjects was conducted [5]. This prospective, cross-sectional, observational study was performed. Thirty (30) patients attending the rheumatoid clinic at one of the major regional hospitals in Trinidad and Tobago were recruited. There was no interventional measure; however, the study sought to observe and delineate the presence of IL-17, 23, and 33 among patients attending a rheumatoid outpatient clinic. Using the Revised American College of Rheumatology criteria, lupus diagnosis was confirmed, and the disease was clinically assessed using the SLE Disease Activity Index (SLEDAI) 2000 score depicted in Table 1 [6]. A score of ≥3 or four was considered an active disease, and a score of <3 was considered an inactive disease. Thirty (30) patients who were non-SLE subjects were also chosen as normal controls for the study.

**Table 1:** **D**escribe SLEDAI 2000 (SLEDAI- 2K), a modification of SLEDAI to reflect persistent, active disease in those descriptors that had previously only considered new or recurrent occurrences; and as a measure of global disease activity in the clinic [6].

| Weight | SLEDAI SCORE | Descriptor | Definition |
| --- | --- | --- | --- |
| 8 | _____ | Seizure | Recent onset, exclude metabolic, infectious or drug causes. |
| 8 | _____ | Psychosis | Altered ability to function in normal activity due to severe disturbance in the perception of reality. Include hallucinations, incoherence, marked loose associations, impoverished thought content, marked illogical thinking, bizarre, disorganized, or catatonic behavior. Exclude uremia and drug causes. |
| 8 | _____ | Organic brain syndrome | Altered mental function with impaired orientation, memory, or other intellectual function, with rapid onset and fluctuating clinical features, inability to sustain attention to environment, plus at least 2 of the following: perceptual disturbance, incoherent speech, insomnia or daytime drowsiness, or increased or decreased psychomotor activity. Exclude metabolic, infectious, or drug causes. |
| 8 | _____ | Visual disturbance | Retinal changes of SLE. Include cytoid bodies, retinal hemorrhages, serous exudate or hemorrhages in the choroid, or optic neuritis. Exclude hypertension, infection, or drug causes. |
| 8 | _____ | Cranial nerve disorder | New onset of sensory or motor neuropathy involving cranial nerves. |
| 8 | _____ | Lupus headache | Severe, persistent headache; may be migrainous, but must be nonresponsive to narcotic analgesia. |
| 8 | _____ | CVA | New onset of cerebrovascular accident(s). Exclude arteriosclerosis. |
| 8 | _____ | Vasculitis | Ulceration, gangrene, tender finger nodules, periungual infarction, splinter hemorrhages, or biopsy or angiogram proof of vasculitis. |
| 4 | _____ | Arthritis | ≥ 2 joints with pain and signs of inflammation (i.e., tenderness, swelling or effusion). |
| 4 | _____ | Myositis | Proximal muscle aching/weakness, associated with elevated creatine phosphokinase/aldolase or electromyogram changes or a biopsy showing myositis. |
| 4 | _____ | Urinary casts | Heme-granular or red blood cell casts. |
| 4 | _____ | Hematuria | >5 red blood cells/high power field. Exclude stone, infection or other cause. |
| 4 | _____ | Proteinuria | >0.5 gram/24 hours |
| 4 | _____ | Pyuria | >5 white blood cells/high power field. Exclude infection. |
| 2 | _____ | Rash | Inflammatory type rash. |
| 2 | _____ | Alopecia | Abnormal, patchy or diffuse loss of hair. |
| 2 | _____ | Mucosal ulcers | Oral or nasal ulcerations. |
| 2 | _____ | Pleurisy | Pleuritic chest pain with pleural rub or effusion, or pleural thickening. |
| 2 | _____ | Pericarditis | Pericardial pain with at least 1 of the following: rub, effusion, or electrocardiogram or echocardiogram confirmation. |
| 2 | _____ | Low complement | Decrease in CH50, C3, or C4 below the lower limit of normal for testing laboratory |
| 2 | _____ | Increased DNA binding | Increased DNA binding by Farr assay above normal range for testing laboratory. |
| 1 | _____ | Fever | >38° C. Exclude infectious cause. |
| 1 | _____ | Thrombocytopenia | <100,000 platelets / x10<sup>9</sup>/L, exclude drug causes. |
| 1 | _____ | Leukopenia | < 3,000 white blood cells / x10<sup>9</sup>/L, exclude drug causes. |
| **TOTAL SLEDAI SCORE** _____ |  |  |  |

The characteristics of the study population included all women aged > 18 years with an active SLE diagnosis. Female patients were enrolled in the study because their prevalence was higher in women than in men. One male patient attending the Rheumatology Clinic for SLE was also recruited in the study, as all available men were included. The target population of this study was 60, of which 30 patients with active SLE disease were included.

### Inclusion and Exclusion Criteria

Only adult patients with active SLE disease and all ethnic groups who gave permission to participate in the project were included.

None of the patients with SLE who refused to participate were included. SLE occurs mostly in women above childbearing age. Although there may be children who could suffer from the disease, they did not fall into this age category as adult females of childbearing age.

Laboratory workup as blood cell count and differential, blood chemistry, C3 and C4 levels, antinuclear antibodies, kidney function test as the presence of proteinuria and haematuria, and other blood routine was done by standard laboratory procedure in the Clinical Laboratory Department at Eric Williams Medical Science complex and were collected from the patient records,

## Methodology

Patients consented to participate in the study. The demographic data of these patients were collected during this period using a standardized questionnaire. Following this, 5 ml of the venous blood sample was drawn from the patient’s arm into red and purple top bottle tubes, and the tubes were taken to the Immunology Laboratory section of the Department of Paraclinical Sciences of the UWI for analysis.

### Immunoassays (ELISA) for the determination of the serum concentration of IL-17, IL-23 or IL-33

The sera were extracted and stored at minus20oC until further analysis. Using commercially available sandwich ELISA kits (IBL International, Hamburg, Germany), the sera were tested to determine the presence of IL-23, 17, and 33 according to established immunochemical analysis and the manufacturer’s instructions.

Three tests were performed using IBL human IL-17A, IL-23, and IL-33 ELISA kits. Ninety-six-well ELISA plates were lined with monoclonal anti-human antibodies against IL-17, IL-23, or IL-33. The patient samples were added to the plates and incubated. After washing, the wells were incubated with biotin-conjugated anti-human 1L 17A, IL 23, or IL 33.

For the IL-17A ELISA kit, and IL-23 ELISA kit protocol followed the guidance as referenced in 5.

For the IL-33 ELISA kit, the wells were incubated with human serum for 1 h, the microplate was washed, and after incubation with the anti-human biotin conjugate for 1 h, the microplate was rewashed, and streptavidin-coupled horseradish peroxidase (HRP) was added. Unbound avidin-HRP was removed by washing, and a reactive substrate solution was added. The levels of IL-33 in the samples were proportional to the amount of colored product produced. The addition of 3M H2SO4 stopped the reaction, and the absorbance of the samples and controls was measured at 450 nm. Human IL-33 concentrations were determined using a standard curve drawn from seven human IL-33 standard dilutions.

### Statistical analysis

Data were analyzed using Microsoft Excel and SPSS 22 software programs. The data were descriptive, and it was reported as a comparison of frequency distributions. A p < 0.05 was considered a statistically significant value. A linear regression test was used to determine the cytokine concentrations in the stated population.

## Results

A total of 60 blood samples were drawn from 30 SLE patients and 30 healthy individuals. The crucial observation from the analysis of the age groups of the patients enrolled in this study, most of the patients who had SLE were in the 25 to 30 years age group. for the controls, and the highest frequency was those over 40 years of age. The least number of participants recruited from the SLE group was under the 20-year age group.

Of all SLE patients, 15 (50%) were of Afro-Trinidadian descent, 12 (40%) were of Indo-Trinidadian descent, and 3 (10%) were of mixed descent or origin. Of all the SLE patients recruited in the investigation, one (3%) was a male patient and 29 (97%) were female. Nineteen healthy controls (63%) were women, and 11 (37%) were men.

These results were related to patient employment and SLE. Most SLE patients were unemployed (47%), while the second most common occupation was a 10% clerk. In non-SLE patients like SLE patients, it was found that many patients were unemployed as well at 57%.

Cytokine concentrations and SLEDAI scores in patients with and without SLE SLEDAI-2K scores ranged from 4 to 14. The mean SLEDAI score was 9.1, and the median score was 9.5. Table 1 shows that many patients (87%) experienced arthritis, and no patients experienced urinary or cardiovascular accidents. Visual disturbances accounted for 35% of patients, while headaches accounted for a significant number of patients at 68%. Vasculitis was seen in 32% of the patients, while renal symptoms such as proteinuria and hematuria accounted for 50% and 30%, respectively. Many patients experienced symptoms of myositis at 77% and alopecia at 60%. Pleurisy accounted for 43% of patients, while low complement and leukopenia values were 60% and 35%, respectively [5-7]. In the control group those manifestations accounted for less that 5%. There was a statistical significance of SLE manifestation percentages as compared with the control group as shown in Table 2.

**Table 2.** Percentage of clinical manifestations based on the Systemic lupus erythematosus (SLE) disease activity index (SLEDAI) 2000 score.

| Clinical Manifestation | Percentage | P-value |
| --- | --- | --- |
| Arthritis | 89% | 0.0000 |
| Visual disturbances | 35% | 0.0026 |
| Headache | 68% | 0.0000 |
| Vasculitis | 32% | 0.0046 |
| Proteinuria | 50% | 0.0001 |
| Haematuria | 30% | 0.0067 |
| Fever | 72% | 0.0000 |
| Myositis | 77% | 0.0000 |
| Pleurisy | 45% | 0.0004 |
| Low complement levels | 60% | 0.0000 |
| Leukopenia | 35% | 0.0026 |

All variables measured are statistically significant, $p < 0.05$ .

Figure 2 shows that serum IL-17 and IL-23 levels were more significant in SLE patients than in controls (P<0.05); however, there was no statistically significant difference between IL-33 levels between SLE patients and healthy controls (P<0.05). There was no significant correlation between serum IL-17 levels and the 23 SLE patients with SLE (r=0.308, p>0.05). Serum IL-17 and IL-23 levels were positively correlated with SLEDAI score. Nevertheless, IL-33 levels showed no correlation with SLEDAI score.

## Discussion

This study aimed to investigate the association between IL-17, 23, and 33 cytokines in SLE. It determines whether there is any correlation between these cytokines and the disease activity index. In a study conducted by Hegab et al., 2014, they looked at IL-23 serum levels in patients with lupus compared to healthy individuals to correlate the serum levels of cytokines with disease activity and its possible role in the pathogenesis of SLE. Serum levels of IL-23 were determined in all patients and controls using a quantitative enzyme-linked immunosorbent assay. They found that serum IL-23 concentration was significantly elevated in SLE patients compared to healthy controls, and it was significantly correlated with the disease activity index [6]. Their report support our findings that serum IL-23 levels were higher in SLE patients than in healthy individuals and positively correlated with the SLEDAI score.

A study conducted by Mok et al., 2010 showed that higher serum IL-17 and IL-23 levels were found in SLE patients than in healthy individuals [3]. Another study conducted by Iwakura et al.,2006 showed that IL-23 induced the differentiation of naive CD4 (+) T cells into pathogenic helper T cells that produce IL-17 and other cytokines vital in the inflammatory response. In his paper, it was also reported that there was a correlation between IL-17 and IL-23 levels [8]. This was inconsistent with our results for the Pearson’s correlation coefficient, which equaled 0.0308, and there was a p-value of 0.017; and these correlations were statistically significant.

Yang et al., 2011 examined whether IL-33 serum levels were associated with lupus. A total of 70 patients with diseases were recruited. Sera were obtained at their clinical visit and compared to the sera from 40 healthy controls. Serum IL-33 levels were significantly higher in patients with SLE than in healthy controls, but were lower than those found in patients with rheumatoid arthritis [4]. These results are different from the results obtained in our study, which found no statistically significant difference between serum IL-33 levels among SLE patients and healthy controls (p>0.05).

Mok et al., 2010 examined the association between serum IL-33 levels in patients with SLE and disease activity. The SLEDAI was used to assess disease activity. Sandwich ELISA measured the IL-33 levels. Elevated serum IL-33 showed similar values among SLE patients and controls and showed no correlation with the SLEDAI score [9], which was similar to our results, which showed no correlation between IL-33 and disease activity index.

Pons-Estel et al., 2010 showed an increased risk among reproductive-age women who were African Americans. Moreover, SLE is two to four times more frequent and more severe among non-white populations worldwide and tends to be more severe in men, the pediatric population, and patients with late-onset lupus [10]. These results were similar to those obtained in our current analysis, where there was an increased incidence of SLE among female patients of reproductive age who were of African origin.

## Conclusion

Cytokines IL-17, IL-23 and IL-33 were measure in patients with SLE in Trinidad and Tobago. A larger study is needed to corroborate the participation of this cytokines in SLE and how positively they correlate with the SLEDAI-2K score. However, using an amount of sample representative of the population it was interested to find in this study that serum IL-17 and IL-23 levels were more significant in SLE patients than in controls, but not IL-33 levels, as well both IL-17 and IL-23 correlated best with the SLEDAI-2K score.

## Data Availability

Data is available upon request but a link does not exist.

---

**Figure 1.**
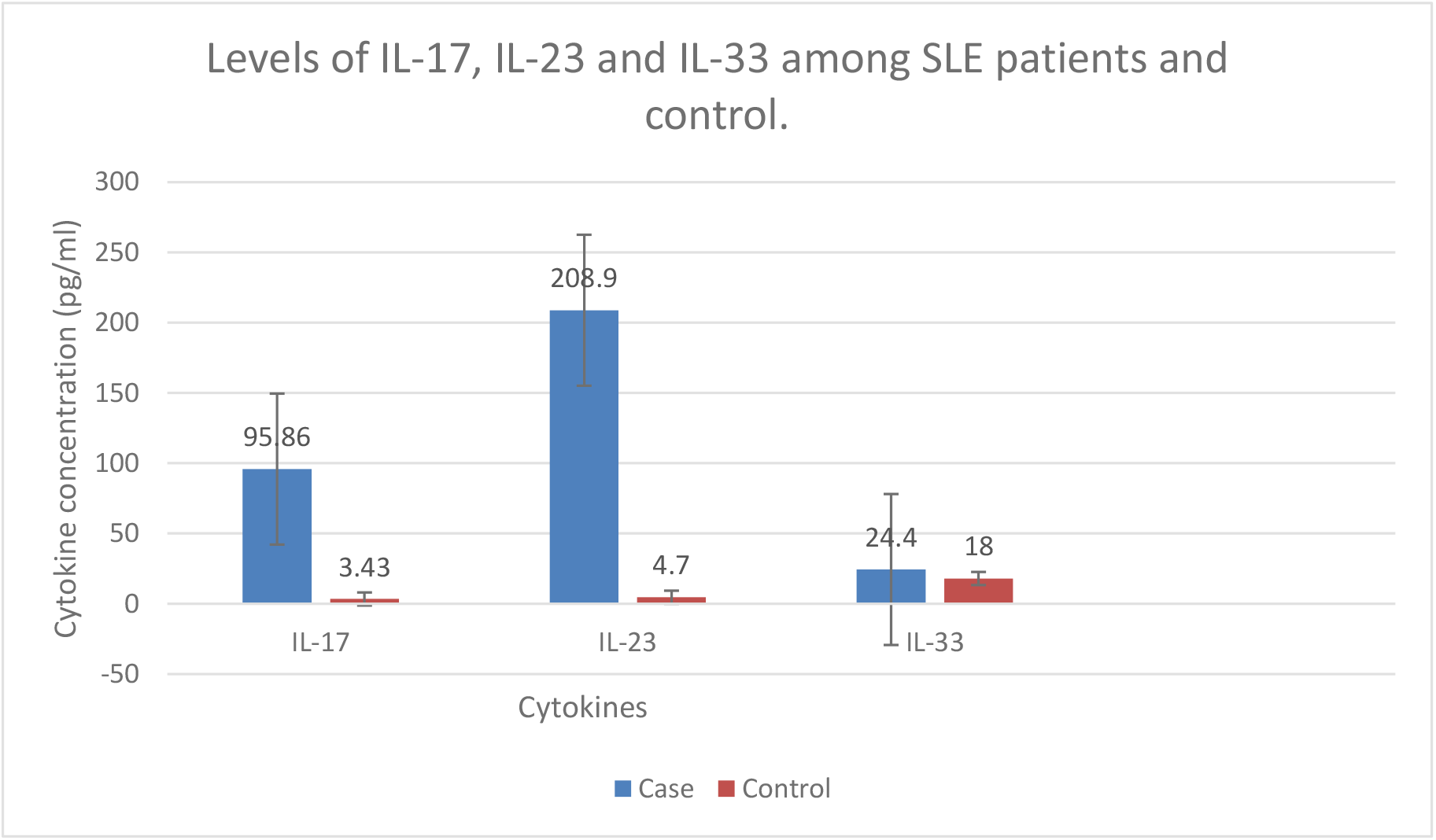
shows the levels of IL-17, IL-23, and IL-33 in patients with and without SLE. The levels of cytokines were significantly higher for IL-23 than for the other ILs. IL-17 levels were significantly higher than in the control group. The control group n=30 was divided into three groups n=10 to compare cytokine levels in patients with SLE patient n=30.

## References

1. Wong CK, Lit LC, Tam LS, Li EK, Wong PT, Lam CW. Hyperproduction of IL-23 and IL-17 in patients with systemic lupus erythematosus: implications for Th17-mediated inflammation in auto-immunity. Clin Immunol 2008;127:385–93.

2. Fabien B, Northcott M., Hoi A, Mackay F, Morand E. Clinical associations of serum IL-17 in systemic lupus erythematosus. Arthritis Research & Therapy 2013; 15:R97.

3. Mok MY, Wu HJ, Lo Y, Lau CS. The relation of IL-17 (IL-17) and IL-23 to Th1/Th2 cytokines and disease activity in systemic lupus erythematosus. J Rheumatol 2010; 37:2046–52.

4. Yang Z, Liang Y, Xi W, Li C, Zhong R. Association of increased serum IL-33 levels with clinical and laboratory characteristics of systemic lupus erythematosus in Chinese population. Clin Exp Med 2011;11:75–80.

5. Nobee A, Justiz-Vaillant A, Akpaka PE, Poon-King P (2016) Levels of Interleukin 17 and 23 in Patients with Systemic Lupus Erythematosus (SLE) in Trinidad and Tobago. Immunochem Immunopathol 2: 115. doi: 10.4172/2469-9756.1000115

6. Gladman DD, Ibañez D, Urowitz MB. Systemic lupus erythematosus disease activity index 2000. J Rheumatol. 2002 Feb;29(2):288-91. PMID: 11838846.

7. Justiz-Vaillant AA, Ferrer-Cosme B, Ramirez-Hernandez N. A Female with Systemic Lupus Erythematosus and Streptococcal Pneumonia treated with Intravenous Immunoglobulins (Ivig). Preprint.org 2020, DOI: 10.20944/preprints202009.0544.v1

8. Iwakura Y, Ishigame H. The IL-23/IL-17 axis in inflammation. J Clin Invest 2006; 116:1218–22.

9. Mok MY, Huang FP, Ip WK, Lo Y, Wong FY, Chan EY, Lam KF, Xu D. Serum levels of IL-33 and soluble ST2 and their association with disease activity in systemic lupus erythematosus. Rheumatology (Oxford) 2010; 49:520–7.

10. Pons-Estel G; Alarcon, Graciela S; Scofield, Lacie Cooper, Glinda S. “Understanding the Epidemiology and Progression of Systemic Lupus Erythematosus”. Seminars in Arthritis and Rheumatism 2010; 39: 257–68.

